# Predictive Model for Severe Coronary Artery Calcification in ESKD Patients

**DOI:** 10.1101/2023.12.16.23300066

**Authors:** Xinfang Tang, Hanyang Qian, Ming Zeng, Hui Huang, Shijiu Lu, Jing Wang, Fan Li, Anning Bian, Xiaoxue Ye, Guang Yang, Kefan Ma, Changying Xing, Yi Xu, Ningning Wang

## Abstract

**Introduction:** The Agatston coronary artery calcification score (CACS) is an assessment index for coronary artery calcification (CAC). This study aims to explore the characteristics of CAC in end-stage kidney disease (ESKD) patients and establish a predictive model to assess the risk of severe CAC in patients.

**Methods:** CACS of ESKD patients was assessed using an electrocardiogram-gated coronary computed tomography (CT) scan with the Agatston scoring method. A predictive nomogram model was established based on stepwise regression. An independent validation cohort comprised of patients with ESKD from multicentres.

**Results:** 369 ESKD patients were enrolled in the training set, and 127 patients were included in the validation set. In the training set, the patients were divided into three subgroups: no calcification (CACS = 0, n = 98), mild calcification (0 < CACS ≤ 400, n = 141) and severe calcification (CACS > 400, n = 130). Among the four coronary branches, the left anterior descending branch (LAD) accounted for the highest proportion of calcification. Stepwise regression analysis showed that age, dialysis vintage, β-receptor blocker, calcium-phosphorus product (Ca × P), and alkaline phosphatase (ALP) level were independent risk factors for severe CAC. A nomogram that predicts the risk of severe CAC in ESKD patients has been internally and externally validated, demonstrating high sensitivity and specificity.

**Conclusion:** CAC is both prevalent and severe in ESKD patients. In the four branches of the coronary arteries, LAD calcification is the most common. Our validated nomogram model, based on clinical risk factors, can help predict the risk of severe coronary calcification in ESKD patients who cannot undergo coronary CT analysis.

## 1. Introduction

The risks of death in chronic kidney disease (CKD) patients are eight times higher than that of the general population, and cardiovascular disease (CVD) accounts for more than 50% of the causes^1^. As an important component of vascular calcification (VC), coronary artery calcification (CAC) is closely related to CVD and high mortality in patients with CKD^2^. In a study of 1,579 participants with CKD G1-G5 without kidney replacement therapy, the results show that the coronary artery calcification score(CACS) was independently associated with adverse cardiovascular outcomes and all-cause death in patients with CKD^3^. Patients with severe coronary calcification whose CACS is greater than 400, have an extremely high atherosclerotic plaque load, which increases their cardiovascular risk^4^. All-cause mortality and cardiovascular events were significantly higher in peritoneal dialysis (PD)patients with a CACS > 400 than in those with a CACS = 0^5^.

In the general population, CACS is evaluated using computed tomography (CT) scans. The Agatston score is valuable for the prognosis of cardiovascular disease^6^. The factors influencing the occurrence and development of CACS include age, body mass index (BMI), diabetes, hypertension, smoking, inflammatory factors, fibrinogen, and dyslipidemia, et^7–11^. Intervention studies on CAC mainly focus on controlling lipid abnormalities^12,13^. In CKD patients, in addition to the above-mentioned influencing factors, several unique pathophysiological characteristics have been observed. These include decreased glomerular filtration rate, hyperphosphatemia, a high calcium-phosphorus product, secondary hyperparathyroidism (SHPT), oxidative stress, systemic inflammation, protein-energy malnutrition, asymmetric dimethylarginine (ADMA), p-cresol, and Fetuin-A. These factors have been proven to be associated with CAC ^5,14–19^. Although CT scanning is a non-invasive imaging method used for detecting CACS, its availablity in primary hospitals is limited.

In this study, we conducted a retrospective analysis of the characteristics of CAC in ESKD patients, using the Agatston CACS. We examined the incidence and clinical features of severe total coronary calcification, defined as a CACS greater than 400, in ESKD patients. The primary objective of this study was to develop a nomogram model that can predict the presence of severe CAC in ESKD patients. The proposed model aims to provide valuable insights for the prevention and treatment of CAC, particularly in primary care settings.

## 2. Materials and Methods

### 2.1. Study Populations

This retrospective study included 369 CKD5 patients who received treatment at the Department of Nephrology, the First Affiliated Hospital of Nanjing Medical University, from June 2017 to April 2022. The patients in this group constituted the training set. The inclusion criteria were as follows^20^: (1) individuals aged between 18 and 75 years old; (2) eGFR (estimated using the CKD-EPI formula) less than 15 ml/min/1.73m^2^; and (3) individuals who had undergone predialysis or regular dialysis for a duration of three or more months.

Exclusion criteria included: (1) Estimated survival time < 6 months; (2) Fever or infection; (3) Pregnant or lactating women; (4) Severe liver disease, chronic obstructive pulmonary disease, malignant tumor or serious mental disease; (5) History of severe congenital heart disease, atrial fibrillation, atrial flutter, high-grade atrioventricular block and permanent pacemaker implantation^21^; (6) Arterial calcification cannot be detected or the results are unreliable, such as arrhythmia, cardiac stent implantation, amputation or serious peripheral vascular diseases^22^; (7) History of other concomitant diseases affecting calcium status in the body and soft tissue calcification^23^, such as sarcomatoid nodules, multiple myeloma, human immunodeficiency virus (HIV, AIDS virus), amyloidosis and primary parathyroid disease.

Additional 127 patients diagnosed with CKD5, who received treatment at Department of Nephrology, the First Affiliated Hospital of Nanjing Medical University and the Affiliated Lianyungang Oriental Hospital of Kangda College of Nanjing Medical University from May 2022 to May 2023, were included as the validation cohort. These patients were selected based on the same criteria as the initial cohort, with the aim of validating the predictive effectiveness of the nomogram model.

### 2.2. Measurements and Assessments

The general information, baseline laboratory examinations, and CACS evaluated using CT scan were documented for all participants. This study was approved by the ethics committee of the First Affiliated Hospital of Nanjing Medical University (ethics No.: 2011-sr-072. 2019-sr-368). All participants have signed informed consent.

#### Research Process

All venous blood samples were collected from participants at 7 a.m. after an overnight fast. These samples were then analyzed for routine blood tests, biochemical indices such as calcium and phosphorus, bone alkaline phosphatase (BAP), 25 hydroxyvitamin D (25-OH-D), and intact parathyroid hormone (iPTH). For hemodialysis(HD) patients, blood samples were obtained prior to dialysis. The assessment of CACS using CT scan must be done within one week of the hematological examination.

#### Laboratory Examinations

The routine blood tests were detected by LH-750 blood cell analyzer (Beckman Coulter, Fullerton, CA, USA). Automatic biochemical analyzer (AU5400; Olympus Corporation, Tokyo, Japan) was used to detect blood biochemical indices. The serum iPTH level was detected by immunoassay system (Unicel Dxi800 Access; Beckman Coulter, Fullerton, CA, USA). The serum BAP level was determined by enzyme-linked immunosorbent assay (ELISA) and serum 25-OH-D level was determined by radioimmunoassay. The correction formula of blood calcium: serum corrected calcium (mmol/l) = total serum calcium (mmol/l) + [40 - blood albumin (g/l)] × 0.02.

#### CACS Evaluation

Continuous sections were obtained without gaps through an electrocardiogram (ECG)-gated coronary CT plain scan with the Agaston soring method. The scan range was from the tracheal carina to 1 cm below the heart apex. All sections were evaluated to determine the presence and number of coronary calcifications. The threshold of calcification was set as a CT density of 130 Hounsfield units (Hu) with an area ≥ 1mm^2^. All pixels with a CT density ≥ 130 Hu were displayed in each section. A "region of interest" was defined around all the calcifications found in the coronary artery to measure the calcification area in mm^2^. The density score was determined based on the maximum CT attenuations in each region of interest automatically: 1 (score) = 130 to 199 HU, 2 = 200 to 299 HU, 3 = 300 to 399 HU, 4 = greater than or equal to 400 HU. The score of each region of interest is calculated by multiplying the density score and the region’s area. The total CACS was determined by adding the scores of all sections.

### 2.3. Statistical Analysis

The continuous variables are presented as mean ± standard deviation or median (Q1, Q3), and the categorical variables are presented as number (percentage). Baseline characteristics between groups were compared using the unpaired, two-tailed t test or Mann-Whitney test for continuous variables, depending on the data distribution, and the χ^2^ test was used for categorical variables.

Univariate logistic regression analysis was used to screen the factors affecting severe LAD calcification (LAD CACS > 183.4) and severe total CACS (total CACS > 400), including the baseline characteristics (age, sex, BMI, blood pressure, dialysis mode, medication history, renal etiology, etc.) and laboratory indicators (hemoglobin (Hb), hematocrit (HCT), glucose, creatinine, urea, total cholesterol (TC), triglyceride (TG), low density lipoprotein cholesterol (LDL-C), high density lipoprotein cholesterol (HDL-C), lipoprotein a (Lpa), serum albumin (Alb), calcium (Ca), corrected Ca, phosphorus (P), calcium-phosphorus product (Ca × P), ALP, BAP, 25-OH-D and iPTH). The results were presented by odds ratio (OR) and 95% confidence interval (CI).

Multivariate logistic stepwise regression analysis was used to analyze the factors related to severe total CACS, including age, BMI, systolic blood pressure, dialysis vintage, diabetes, medication history (including lipid-lowering treatment, ACEI/ARB, β-receptor blocker, and cinacalcet) and laboratory indicators (including TC, TG, LDL-C, HDL-C, Ca × P, ALP and BAP).

According to the results of multiple stepwise regression analysis, a nomogram model was established to predict the risk of severe CACS in ESKD patients, and used the area under the curve (AUC) value of the receiver operating characteristic (ROC) curve to evaluate the predictive performance of the model. In addition, we conducted external validation in an independent case-control study (n = 127).

All statistical analyses were performed using R foundation for Statistical Computing (version 4.3.1), unless otherwise specified. A P*-*value of < 0.05 was considered statistically significant.

## 3. Results

### 3.1. General Information of the Study Population

A total of 369 ESKD patients were enrolled, including predialysis patients (13.01%), HD patients (64.23%), and PD patients (23.76%). The average age of participants was 49.41 ± 12.14 years old, and the average dialysis vintage was 73.00(24.00-120.00) months. The drugs used to treat chronic kidney disease-mineral and bone disorders (CKD-MBD) include phosphorus binders (39.57%), active vitamin D sterols (35.50%), and cinacalcet (27.10%).

The patients were divided into three subgroups according to the overall degree of CACS: no calcification (CACS = 0, n = 98), mild calcification (0 < CACS ≤ 400, n = 141), and severe calcification (CACS > 400, n = 130). Age, BMI, diastolic blood pressure, dialysis mode, dialysis vintage, whether complicated with hypertension, and the use of β-receptor blockers were significantly different among the subgroups (*P* < 0.05). Among these, haemodialysis patients (43.46%) are more likely to have severe CAC compared to peritoneal dialysis patients (22.36%). The serum Hb, HCT, TC, LDL-C, HDL-C, Ca, corrected Ca, Ca × P, log(ALP), log(BAP), and log(iPTH) also showed significantly different (*P* < 0.05) (Table 1).

**Table 1.**
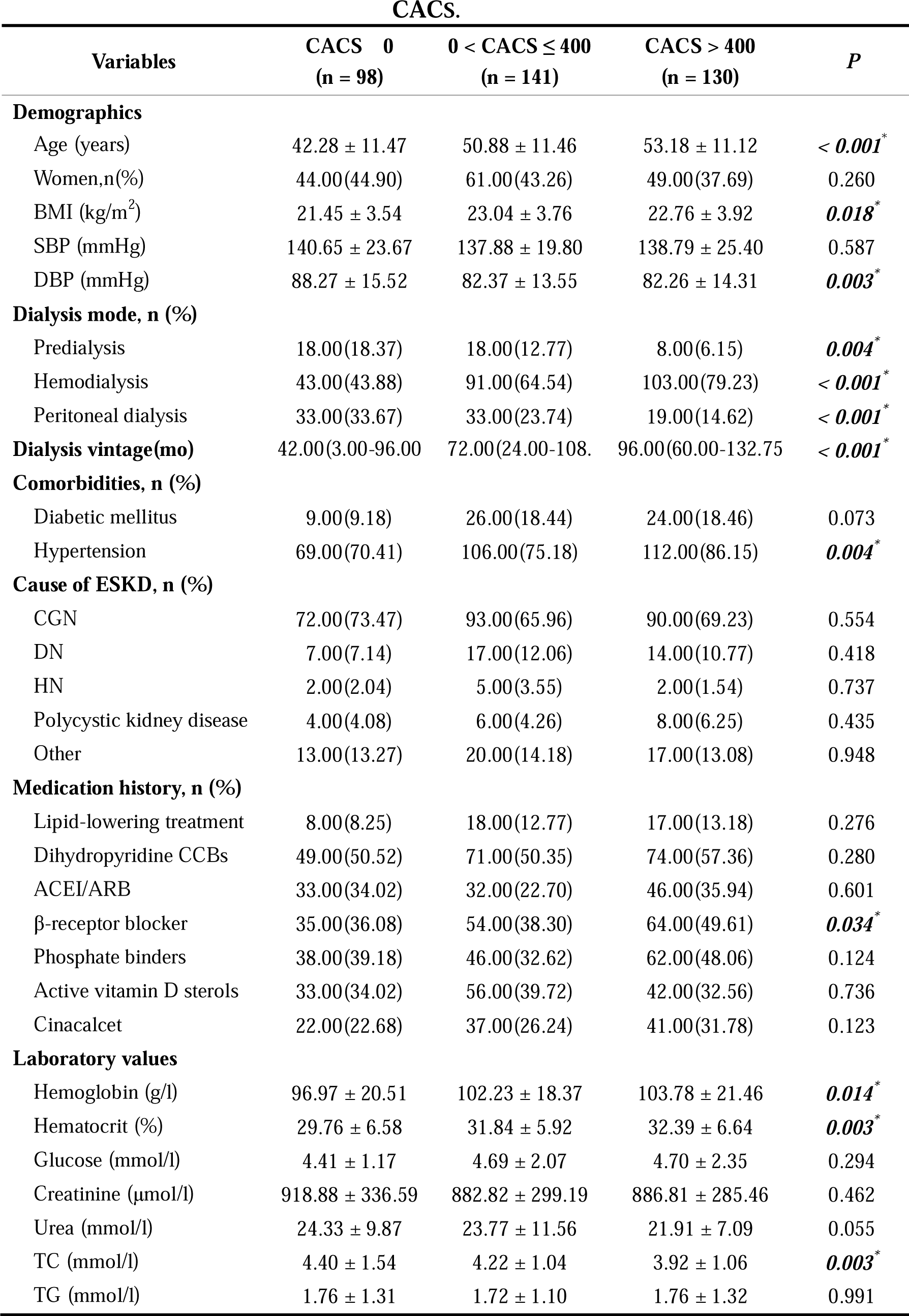
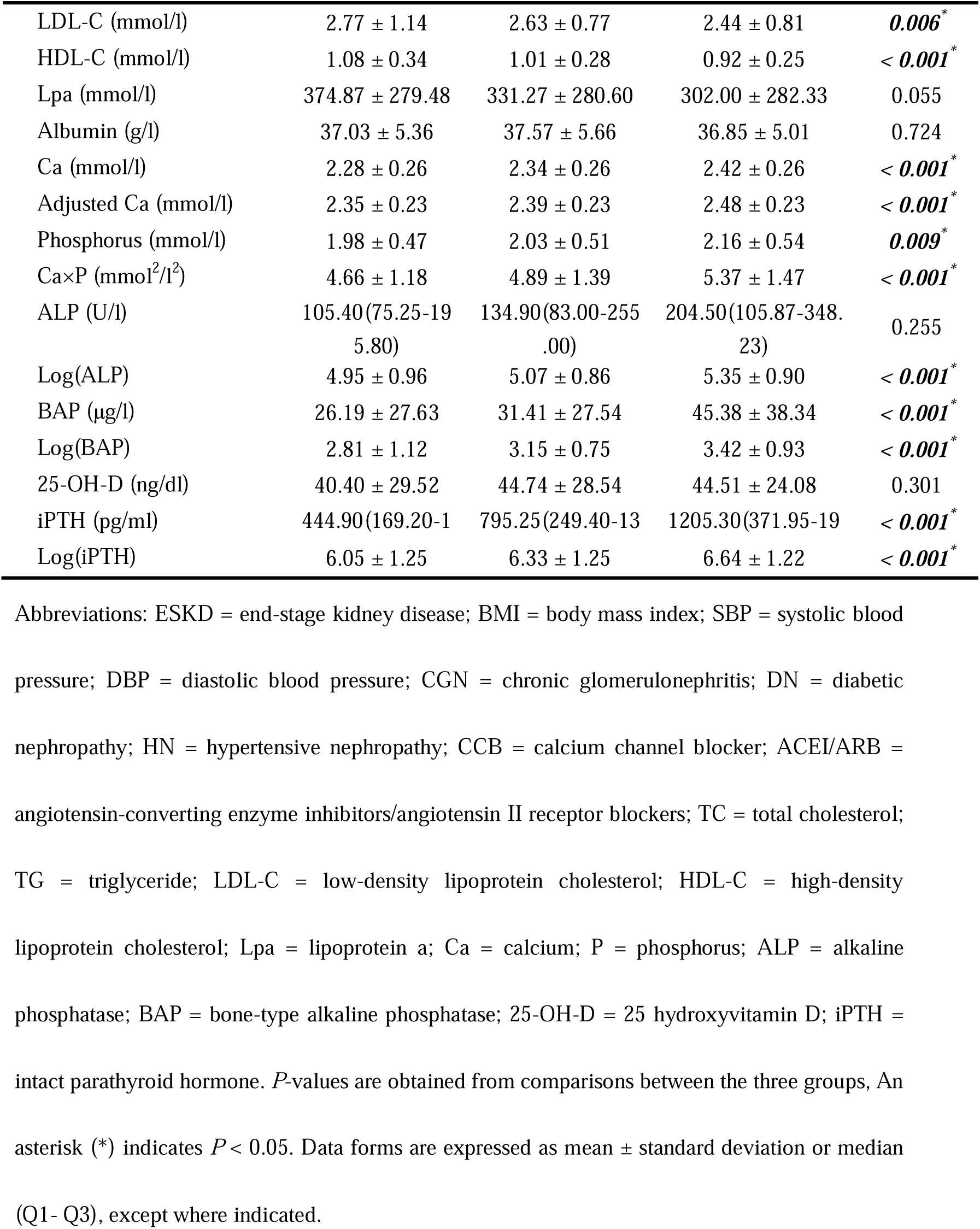
Clinical Characteristics and Laboratory Results of ESKD Patients Subgrouped by CACS.

### 3.2. The Percentage of LAD Calcification was Highest Among the Four Coronary Branches in ESKD Patients

We conducted an observation of calcification in four coronary branches (Figure 1). Out of the 369 ESKD patients, 248 individuals (67.21%) exhibited calcification in LAD, 85 individuals (23.04%) exhibited calcification in the left main trunk (LM) calcification, 175 individuals (47.42%) exhibited calcification in the circumflex branch (CX), and 204 individuals (55.28%) exhibited calcification in the right coronary artery (RCA) (*P* < 0.05). The percentage of LAD calcification was highest among the four coronary branches in ESKD patients.

**Figure 1.**
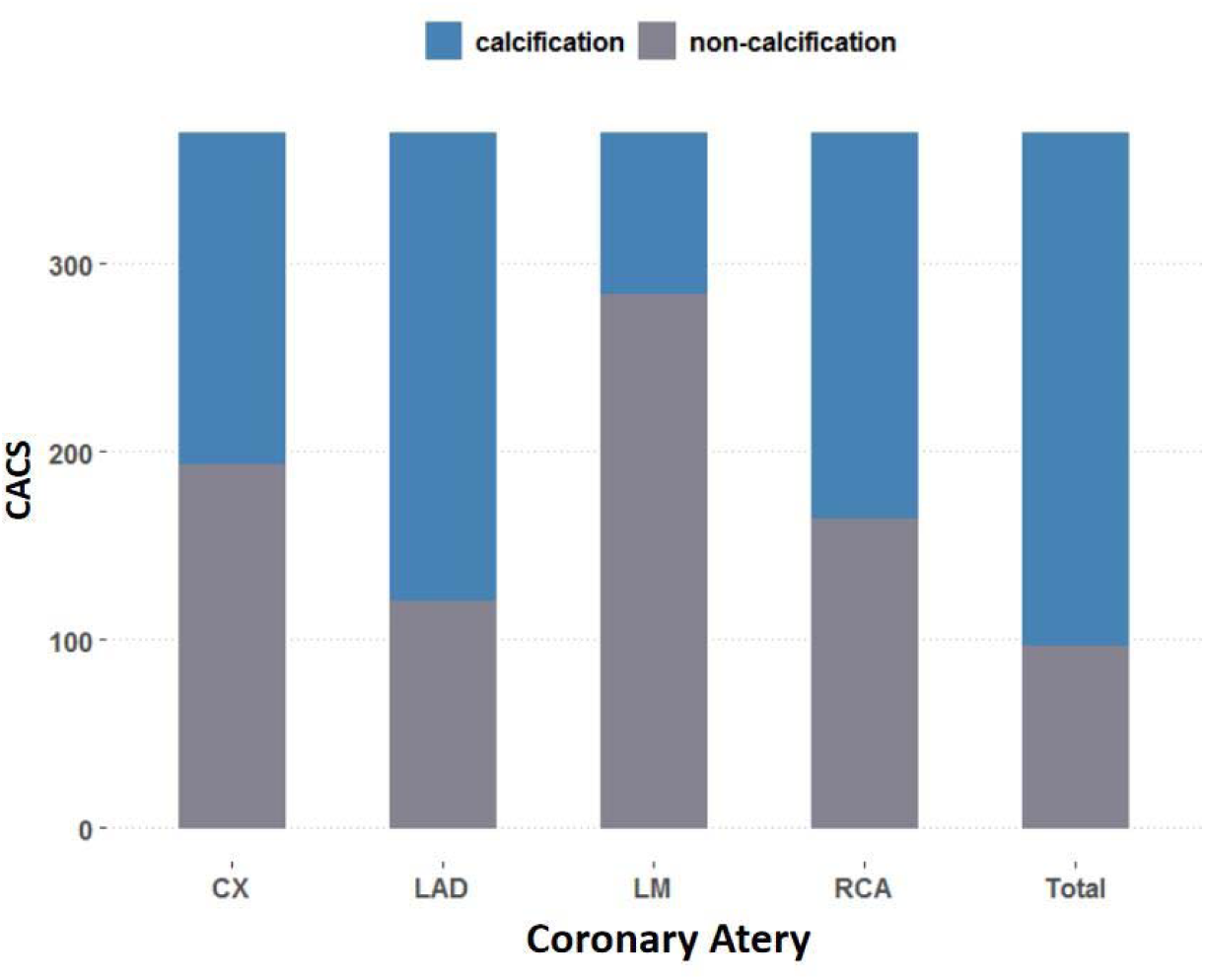
Comparison of Coronary Branches Calcification in Patients with ESKD. Abbreviations: LAD = left anterior descending branch; LM = left main trunk; CX = circumflex branch; RCA = right coronary artery; CACS = coronary artery calcification score.

### 3.3. Imaging Characteristics of ESKD Patients with Varying Degrees of CAC

Figure 2 depicts CT images of patients exhibiting different degrees of CAC. Specifically, Figure 2a illustrates a patient who does not present coronary calcification (calcification score: 0). In contrast, Figure 2b displays a patient with sole calcification in the LAD (calcification score: LAD = 647.9). Figure 2c (calcification score: LM = 156.5, LAD = 2320.7) and 2d (calcification score: CX = 2317.5, RCA = 2329.3) show calcification of 4 coronary branches in one patient.Figure 2c displays the calcification score for the LM as 156.5, and the LAD as 2320.7. Figure 2d, the calcification score for the CX is 2317.5, and for the RCA is 2329.3. Figure 2c and Figure 2d collectively illustrate the calcification of all four coronary branches in a single patient.

**Figure 2.**
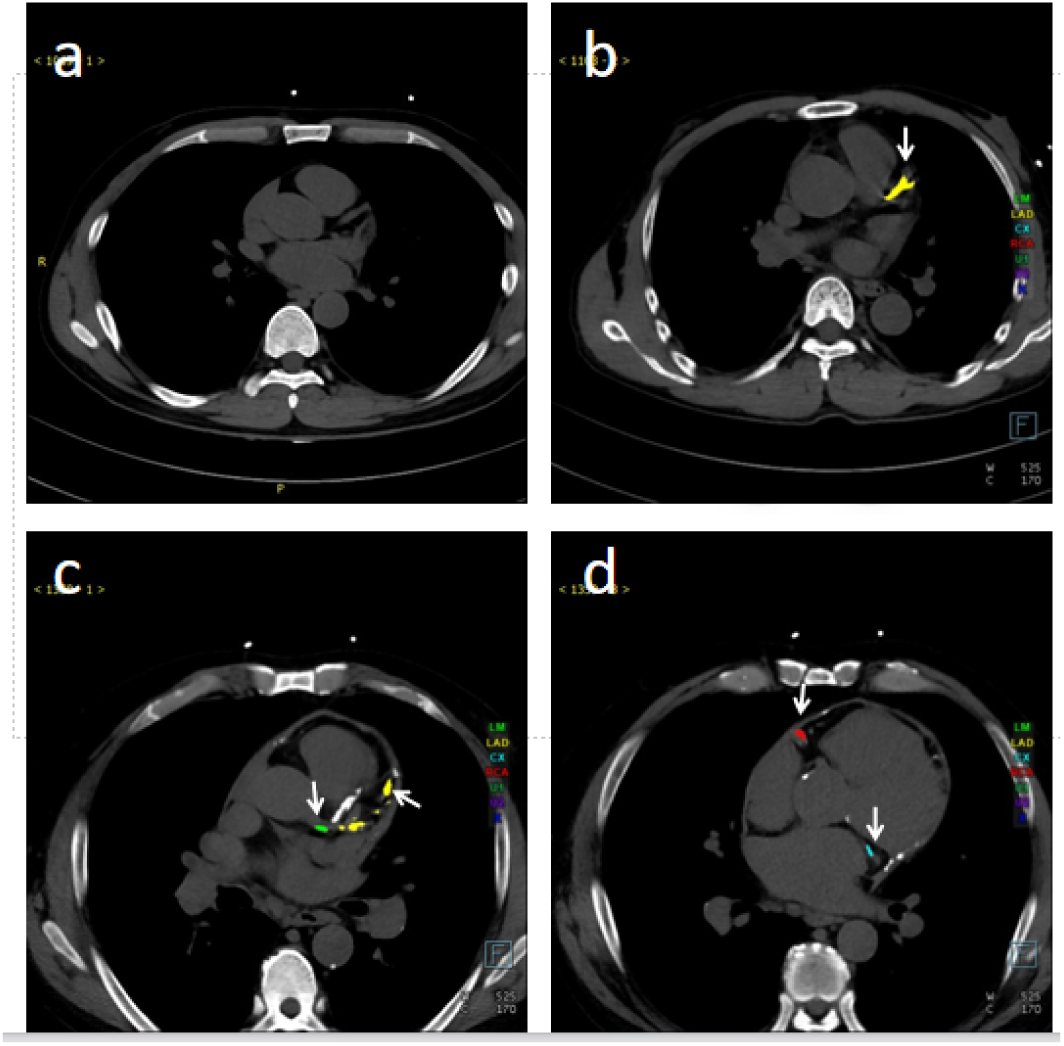
Cross-sectional CT Images of ESKD Patients with Varying Degrees of CACS. (a) An ESKD patient without coronary calcification. (b) An ESKD patient with calcification only in the LAD. (c-d) An ESKD patient with calcification in all four coronary branches. (c) Calcification in the left main stem and left anterior descending branch. (d) Calcification in the circumflex branch and right coronary artery. Different colors represent each coronary branch with calcification (green: left main trunk; yellow: left anterior descending branch; blue: circumflex branch; red: right coronary artery), and the arrows indicated the calcified branches. Abbreviations: CT = computed tomography; ESKD = end-stage kidney disease; LAD = left anterior descending branch.

### 3.4. Clinical Characteristics of LAD Calcification in ESKD Patients

A total of 369 patients were categorized into three subgroups based on their LAD calcification score namely: low (score of 0), intermediate (score ranging from 0 to 183.4), and high (score exceeding 183.4) (Table S1). The clinical characteristics and laboratory results of these subgroups were observed and analyzed. Statistical analysis revealed significant differences in age, BMI, systolic blood pressure, dialysis mode, dialysis vintage, and the presence of concomitant hypertension among the three subgroups (*P* < 0.05). There were statistically significant differences observed in the laboratory indicators between the subgroups, including Hb, HCT, TC, HDL-C, LDL-C, Ca, adjusted Ca, P, Ca × P, log(ALP), log(BAP), and log(iPTH) (*P* < 0.05).

### 3.5. Risk Factors Associated with Severe CAC in Patients with ESKD

We examined the risk factors associated with severe calcification of the LAD, as indicated by a LAD coronary artery calcium score (CACS) exceeding 183.4, as well as the overall CACS exceeding 400 (Table S2). Through univariate analysis, we observed that age, dialysis mode, dialysis vintage, and the use of β-blockers demonstrated significant associations with the occurrence of severe CAC in ESKD patients (*P* < 0.05). Patients with HD were found to have a higher propensity for developing severe CAC compared to patients with PD (odds ratio [OR] 2.989; 95% confidence interval[CI] 1.823-4.903; *P* < 0.001); in contrast, patients with PD exhibited a lower incidence of severe CAC (OR 0.444; 95% CI 0.253-0.779; *P* < 0.05). Further analysis revealed significant correlations between blood lipid metabolism-related indicators, specifically TC, LDL-C, and HDL-C levels, and the development of severe CAC. Blood bone metabolic indexes, including Ca, adjusted Ca, P, Ca × P, log(ALP), log(BAP) and log(iPTH) levels, were observed to have a statistically significant association with the risk of severe CAC (*P* < 0.05). The risk factors related to severe calcification of the LAD showed similar consistency with those associated with total CACS, with the exception of concomitant hypertension and the use of oral phosphorus binders, which were not found to be linked to severe LAD calcification.

### 3.6. Stepwise Regression Analysis of Risk Factors Associated with Severe CAC in Patients with ESKD

A stepwise regression analysis was conducted to identify the risk factors associated with severe CAC in patients with ESKD (Table 2). After adjusting for confounding variables, two independent risk factors for severe CAC in ESKD patients were identified: age (OR, 1.071; 95% CI, 1.037-1.106; *P* < 0.001) and dialysis vintage (OR,1.008; 95% CI, 1.001-1.015; *P* = 0.028). Additionally, the use of oral β-receptor blocker was found to be an independent risk factor for the development of severe CAC (OR, 2.309; 95% CI, 1.095-4.870; *P* = 0.028). Among the blood bone metabolic indexes examined, only Ca×P (OR, 1.333; 95% CI, 1.007-1.771; *P* = 0.045) and log(ALP) (OR, 3.026; 95% CI, 1.080-8.484; *P* = 0.035) levels exhibited a significant association with the occurrence of severe CAC .

**Table 2.**
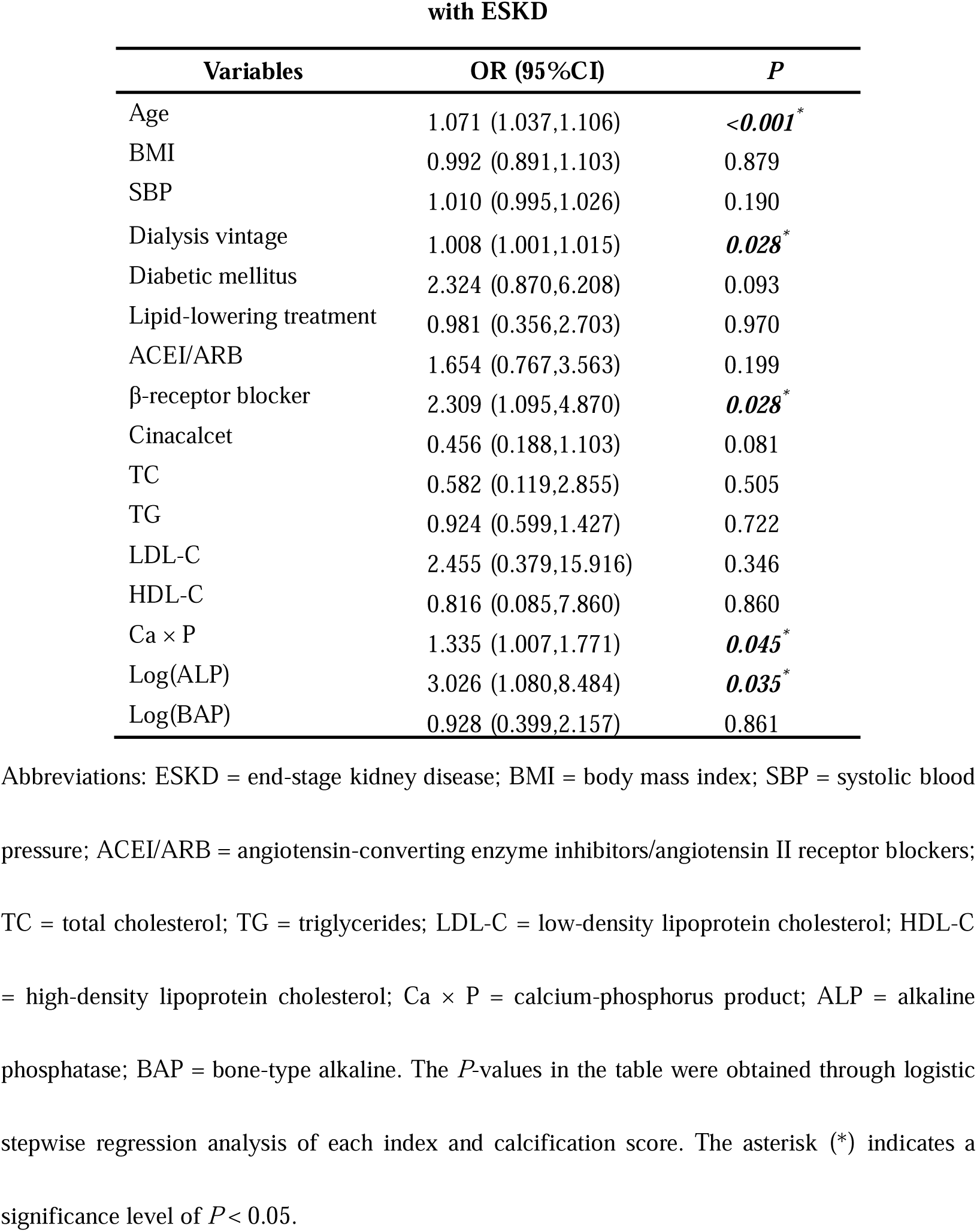
Stepwise Regression Analysis of Clinical Data and Severe Total CACS in Patients with ESKD.

### 3.7. Nomogram Model Based on Clinical Data for Predicting the Risks of Severe CAC in ESKD Patients and Internal Validation

In order to predict the risks of severe CAC in ESKD patients, a nomogram model was established. This model takes into account various clinical factors, including age, BMI, SBP, dialysis vintage, history of diabetes, medication history, and laboratory indicators such as TC, TG, LDL-C, HDL-C, Ca × P, log(ALP), and log(BAP) (Figure 3).

**Figure 3.**
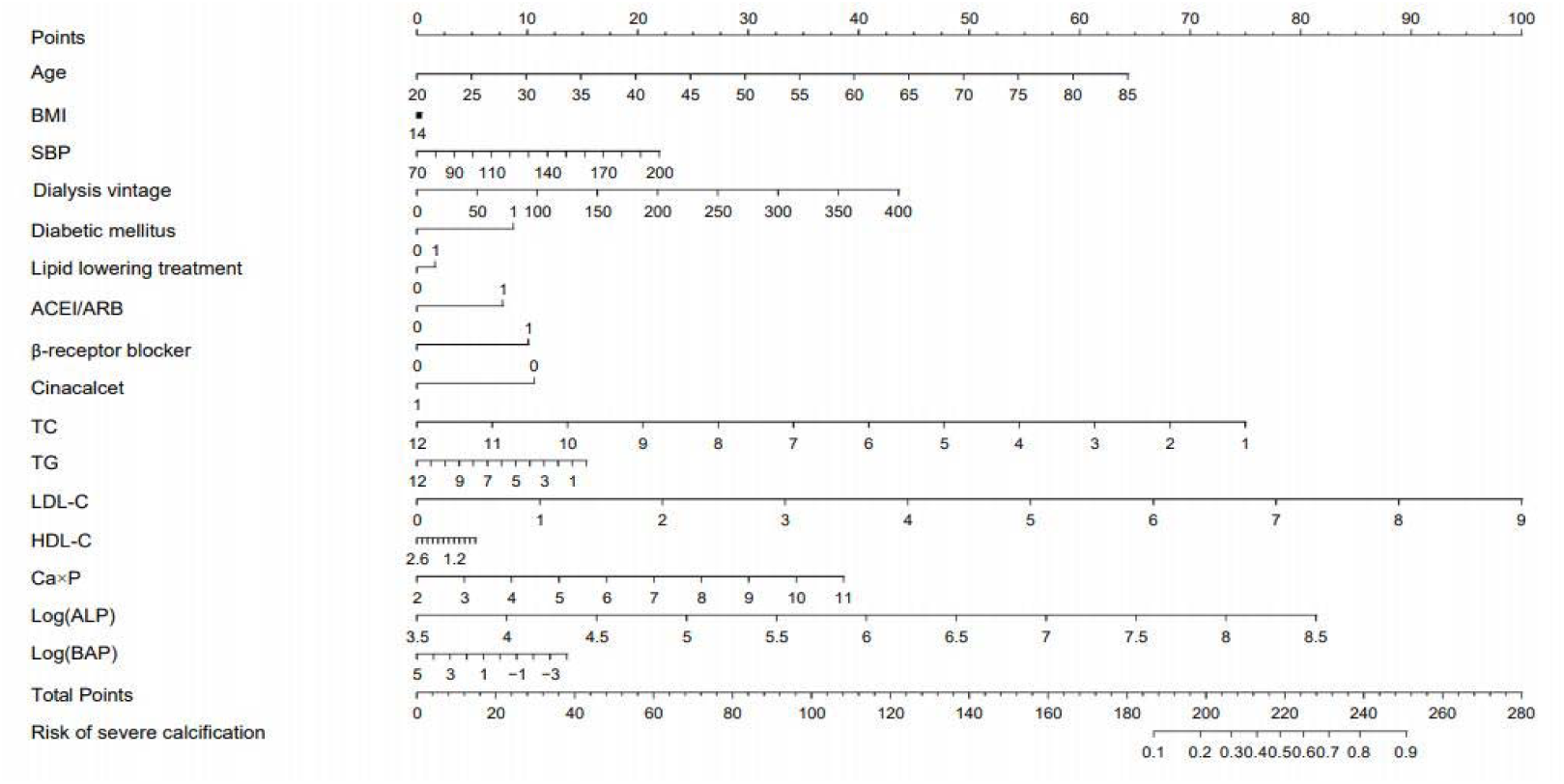
Nomogram Model to Predict the Risks of Severe CAC Based on Clinical Data of ESKD Patients. Based on the value assigned to each patient indicator (0 or 1 for diabetes history and medication history), a dot was plotted on the horizontal line corresponding to the respective item. A vertical line was then drawn upwards to intersect the horizontal line representing the score, thus determining the specific score for that particular item. The total score was calculated by adding up the scores from each item. A dot was plotted on the horizontal line representing the total score. A vertical line was then drawn downwards to intersect the horizontal line corresponding to the risk of severe coronary calcification, giving us the specific value for calcification risk.

The nomogram model’s predictive performance was internally validated using AUC analysis of the ROC curve (Figure 4a). The AUC of this model was determined to be 0.808 (95% CI: 0.751-0.866). Additionally, the sensitivity and specificity were calculated to be 0.843 (95% CI: 0.758-0.928) and 0.701 (95% CI: 0.629-0.772), respectively. These findings suggest that the histogram based on clinical data exhibits a reliable predictive ability in estimating the risks of severe CAC in ESKD patients, demonstrated by its high sensitivity and specificity.

**Figure 4.**
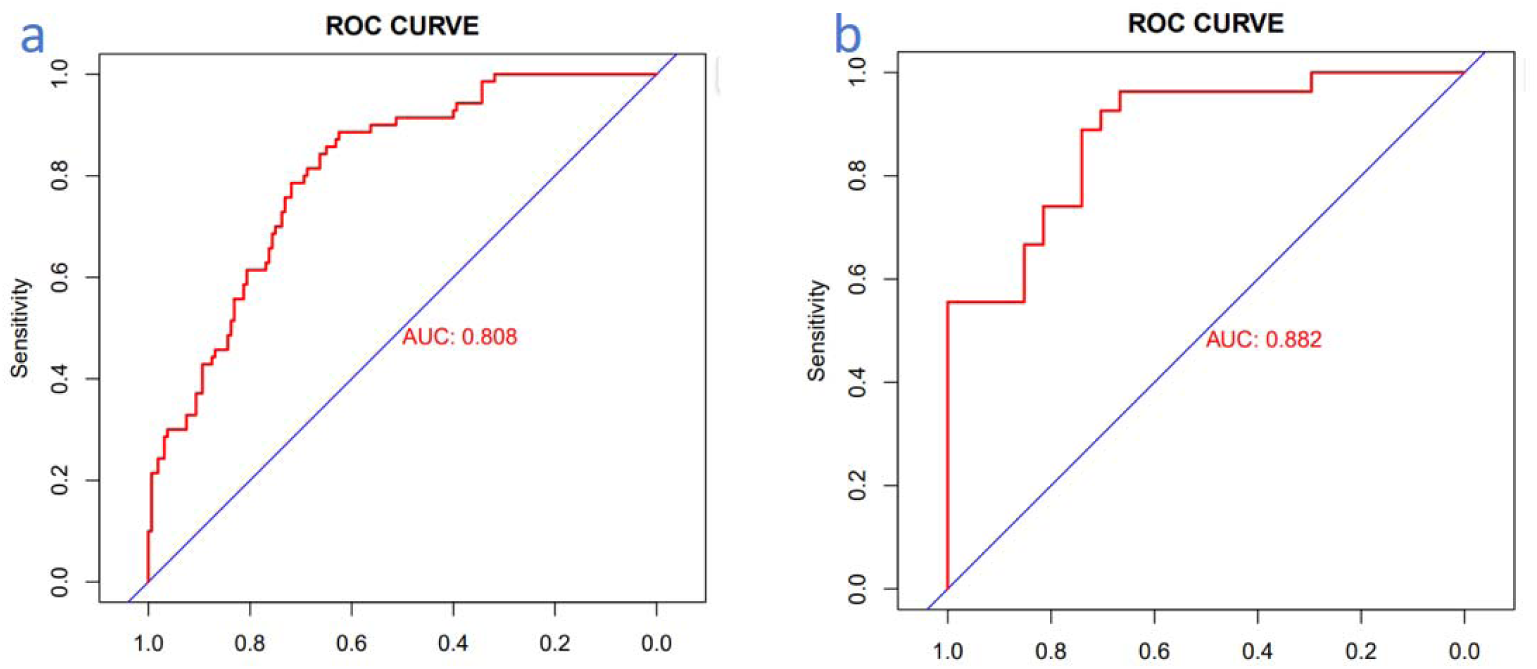
ROC Curves of Nomogram Model. **(a) ROC curve of the nomogram in the training cohort; (b) ROC curve of the nomogram in the validation cohort.** Note: The red curve represents the model’s performance in predicting the risks of severe coronary calcification, with the horizontal axis representing specificity and the vertical axis representing sensitivity. Abbreviations: AUC = area under curve; ROC = receiver operating characteristic.

### 3.8. Independent Validation of the Nomogram

A validation set consisting of 128 patients with stage 5 CKD were utilized for independent validation of the nomogram. This set comprised patients categorized into three groups based on their coronary calcification levels: those with no coronary calcification (n = 22, 17.32%), individuals with mild coronary calcification (n = 51, 40.15%), and subjects with severe coronary calcification (n = 54, 42.52%). Among the patients, 80 (63.00%) were males and 47 (37.00%) were females. The mean age of the patients was 52.41 ± 14.34 years, with a mean duration of dialysis of 60.00 (24.00-124.00) months. The average BMI was 23.10 ± 3.89 kg/m^2^. In terms of comorbidities, 19.69% of the patients with ESKD had diabetes mellitus, and 81.10% of the patients had hypertension (Table S3).

The predictive model underwent external validation through AUC analysis of the ROC curve, utilizing an independent validation dataset (Figure 4b). The model achieved an AUC score of 0.882 (95% CI: 0.794-0.970), with a sensitivity of 0.926 (95% CI: 0.827 -1.000) and a specificity of 0.704 (95% CI: 0.531-0.876). These findings suggest that the predictive model demonstrates commendable predictive capabilities.

## 4. Discussion

In the Multi-Ethnic Study of Atherosclerosis (MESA) project^24^, 684 participants with a baseline eGFR < 60 ml/min/1.73m^2^ had a 66% incidence rate of CAC. Przemyslaw reported that the proportion of CAC in HD patients was 73.1%, which was higher than that in the control group (35.7%) ^19^. After following up with 179 PD patients for 30.6 ± 16.2 months and 104 HD patients for 43.8 ± 19.3 months^25^, CACS was proven to be an independent predictor of all-cause mortality, CVD, and cardiovascular mortality in ESKD patients. In our study, 73.44% of ESKD patients had CAC, and 35.23% had severe calcification (CACS > 400). Previous studies have reported that the severity of CAC is more significant in ESKD patients with a longer duration of dialysis^17,26^, which aligns with our findings. We are the first to reveal that HD patients are more likely to develop severe CAC compared to PD patients.

CACS in CKD patients has been found to correlate with various traditional cardiovascular risk factors, including age, gender, BMI, SBP, diabetes, and use of antihypertensive medications^14,26,27^. In our study, we observed differences in age, BMI, DBP, and hypertension among subgroups stratified based on the severity of CAC. Upon conducting a multivariate regression analysis, age emerged as an independent risk factor for severe CAC, which is onsistent with previous research conducted at our centre^28^. Among the four main coronary branches, the LAD is the most commonly affected vessel in cardiovascular disease^29^, and exhibits the highest proportion of calcification, accounting for 67.21%. We have demonstrated that the factors influencing LAD calcification are comparable to those affecting the total CACS.

Our study revealed that levels of serum-corrected calcium and phosphorus were identified as risk factors for severe CAC. Elevated serum calcium levels were found to directly influence the progression of CAC^30^. Furthermore, hyperphosphatemia was shown to be positively associated with CAC in patients with CKD^21^. Consistently strict phosphate control may slow the progression of coronary and valvular calcifications in incident patients undergoing hemodialysis^31^. In our study, we found that the Ca × P product was an independent risk factor for severe CAC. Additionally, elevated levels of ALP were found to promote vascular calcification, and a positive correlation was observed between high ALP levels and CACS^32^. In a previous study, we also observed a significant association between serum ALP levels and cardiac valve calcification in maintenance HD patients^33^. In line with these findings, we discovered that the ALP served as an independent risk factor for severe CAC. BAP, known as the most important marker of osteoblast differentiation, was reported to be a risk factor for coronary calcification in male HD patients^34^. Finally, serum iPTH levels were positively correlated with CACS^35^, confirming our own finding.

In our study, there was no discernible association between oral cinacalcet, serum 25-OH-D levels, and severe coronary artery calcification. Previous research suggests that Vitamin D deficiency is linked to considerable vascular calcification in CKD patients^36^. Nevertheless, a study involving older African American women revealed no significant relationship between abdominal aorta and serum 25-OH-D levels^37^. A large cross-sectional study found a U-shaped relationship between serum 25(OH)D concentration and the risk of abdominal aortic calcifcation(AAC) and severe AAC^38^. Although it has been established that the combination of cinacalcet and low-dose vitamin D can alleviate CAC^16^, another prospective study involving HD patients with SHPT^39^ found no significant difference between the effect of cinacalcet combined with standard therapy versus standard therapy alone on vascular calcification.

In this study, we find the association of oral β-receptor blockers with the development of severe CAC. Past research has established the presence of β2-adrenergic receptor (AR) on the surface of human osteoblasts (OBs), and it has been suggested that β2-AR agonists could potentially hinder the proliferation of OBs^40^. Wu et al.^41^ discovered that the β-receptor blocker propranolol increased the expression levels of osteogenesis-associated genes, such as bone morphogenetic protein (BMP2), RUNX family transcription factor (RunX2), collagen (COL-1), and osteocalcin (OCN). Consequently, this augmentation promotes the osteogenic differentiation of mesenchymal stem cells (MScs) and OBs. Vascular calcification is a gene-regulated biological process similar to bone mineralization, involving osteogenic differentiation^42^. However, further investigations are necessary to better understand the effects of β-receptor blockers on CAC.

The TC concentration has been proven to be independently associated with the incidence rate of CAC. Hypercholesterolemia is considered a risk factor for CAC^43^. There is a positive correlation between LDL-C and CACS^13,44^. LDL-C levels have also been positively correlated with the risk of CAC events, although this association diminishes with increasing HDL-C levels^13^. In contrast, HDL-C has been shown to be negatively correlated with CACS^27^. However, one study reported that HDL-C and its sub-components (HDL2-C and HDL3-C), as well as the HDL2-C/HDL3-C ratio, had no significant relation with the presence or degree of CAC^45^. High TG levels are related to the progression of CAC^44,46^. Lpa has been established as an independent risk factor for CAC^47,48^. In a study involving the China Han population without diabetes and hypertension, Lpa exhibited a positive correlation with CACS^49^. However, in a study conducted on individuals of African descent did not find an independent correlation between Lpa and CACS^50^. . In our study, we found no correlation between lipid metabolism indices and severe CACS.

Here, we developed a predictive nomogram model to predict the risk of CACS > 400 in ESKD patients. The nomogram model takes into account various factors such as patients’ age, BMI, SBP, dialysis duration, diabetes, medication history, and laboratory examination results, including TC, TG, LDL-C, HDL-C, Ca × P, ALP, and BAP. Both internal and external validation, as indicated by the AUC of the ROC curve, demonstrated the excellent sensitivity and specificity of this nomogram model. Given the challenges associated with conducting CT evaluations for CACS in primary hospitals, the nomogram model serves as a valuable clinical tool for assessing the risks of severe CAC in ESKD patients. It offers a simplified and user-friendly approach that enhances usability in clinical settings.

There are several limitations in our research. This is a retrospective and observational study, lacking relevant bone metabolism markers such as Klotho and FGF23 on haematological examination. The possibility of racial and regional bias cannot be ruled out. The sample size is small, and there is a lack of intervention and follow-up analysis.

## 5. Conclusion

CAC is both common and severe in ESKD patients. Those on HD patients are more susceptible to severe CAC compared to those on PD. Notably, the percentage of LAD calcification is the highest among the four coronary branches in ESKD patients. Various factors have been identified as independent risk factors for severe CAC, including age, dialysis vintage, use of oral β-receptor blocker, serum Ca × P levels, and ALP levels. Formulating a nomogram model based on clinical data can facilitate the prediction of severe CAC risks in ESKD patients. This can contribute to a reducion in cardiovascular events and mortality rates by improving accurate diagnosis and treatment capability, especially for CKD-MBD patients in primary healthcare settings.

## Disclosure

The authors have no conflicts of interest to declare.

## Data Availability

The data that support the findings of this study are available from the corresponding author, upon reasonable request.

## Acknowledgements

This work was funded by the National Natural Science Foundation of China (81270408, 81570666), International Society of Nephrology (ISN) Clinical Research Program (18-01-0247), Jiangsu Provincial Special Program of Medical Science (BL2014080), Construction Program of Jiangsu Provincial Clinical Research Center Support System (BL2014084), Chinese Society of Nephrology (13030300415), Jiangsu Province Key Medical Personnel Project (RC201162, ZDRCA2016002), Six Major Talents Summit of Jiangsu Province (2010-WS-026). The authors thank the affected individuals and their families for participation in this study. The authors thank all nephrologists who referred samples from patients.

## Author Contributions

N.W. and Y.X. devised the conceptual ideas. N.W., Y.X., X.T., and H.Q. contributed to the study design. M.Z., X.T., H.Q., S.L., F.L., and X.Y., enrolled medical records and entered data. M.Z., X.T., H.Q., J.W., F.L., A. B., G.Y., and X.Y. were responsible for the management of stage 5 chronic kidney disease patients. Y.X. and K.M. were in charge of detection of CACS for ESKD patients. X.T., H.Q. and H.H. carried out the statistical analyses and reported the results. N.W., X.T., H.Q., H.H., M.Z, and Y.X. contributed to manuscript writing. All authors reviewed the manuscript and signed off on its accuracy.

## Supplementary Material

Table S1. Clinical Characteristics and Laboratory Results of LAD Calcification in ESKD Patients.

Table S2. Univariate Regression Analysis of Clinical Data and Severe CACS in Patients with ESKD.

Table S3. Clinical Characteristics and Laboratory Results of ESKD Patients Subgrouped by CACS in the Validation Cohort.

