## supplemental tables for "Predictive Model for Severe Coronary Artery Calcification in ESKD Patients"

### ^1^Department of Nephrology, the First Affiliated Hospital with Nanjing Medical University, Jiangsu Province Hospital, Nanjing, China

### ^2^Department of Nephrology, the Affiliated Lianyungang Oriental Hospital of Kangda College of Nanjing Medical University, Lianyungang, China

^3^Department of Nephrology, Nanjing Tongren Hospital, Nanjing, China

^4^Center for Medical Big Data, Nanjing Drum Tower Hospital, Affiliated Drum Tower Hospital, Medical School of Nanjing University, Nanjing, China

^5^Department of Nephrology,Nanjing BenQ medical center, Nanjing, China

^6^Department of Critical Medicine, Geriatric Hospital of Nanjing Medical University, Nanjing, China

^7^Department of Imaging, the First Affiliated Hospital with Nanjing Medical University, Jiangsu Province Hospital, Nanjing, China

^△^These authors contributed equally to this work.

^*^Corresponding Author

Ningning Wang

Department of Nephrology, the First Affiliated Hospital with Nanjing Medical University

Guangzhou Road 300, Nanjing, Jiangsu, 210029, China

Yi Xu

Department of Imaging,the First Affiliated Hospital with Nanjing Medical University,

Guangzhou Road 300, Nanjing, Jiangsu, 210029, China

**KEY WORDS**: End-stage kidney disease; Coronary artery calcification; Agatston coronary artery calcification score; Left anterior descending artery; Chronic kidney disease-Mineral and Bone Disorders; Nomogram

**Abstract**

**Introduction**: The Agatston coronary artery calcification score (CACS) is an assessment index for coronary artery calcification (CAC). This study aims to explore the characteristics of CAC in end-stage kidney disease (ESKD) patients and establish a predictive model to assess the risk of severe CAC in patients.

**Methods**: CACS of ESKD patients was assessed using an electrocardiogram-gated coronary computed tomography (CT) scan with the Agatston scoring method. A predictive nomogram model was established based on stepwise regression. An independent validation cohort comprised of patients with ESKD from multicentres.

**Results**: 369 ESKD patients were enrolled in the training set, and 127 patients were included in the validation set. In the training set, the patients were divided into three subgroups: no calcification (CACS = 0, n = 98), mild calcification (0 < CACS **≤** 400, n = 141) and severe calcification (CACS > 400, n = 130). Among the four coronary branches, the left anterior descending branch (LAD) accounted for the highest proportion of calcification. Stepwise regression analysis showed that age, dialysis vintage, β-receptor blocker, calcium-phosphorus product (Ca × P), and alkaline phosphatase (ALP) level were independent risk factors for severe CAC. A nomogram that predicts the risk of severe CAC in ESKD patients has been internally and externally validated, demonstrating high sensitivity and specificity.

**Conclusion**: CAC is both prevalent and severe in ESKD patients. In the four branches of the coronary arteries, LAD calcification is the most common. Our validated nomogram model, based on clinical risk factors, can help predict the risk of severe coronary calcification in ESKD patients who cannot undergo coronary CT analysis.

**Table S1. Clinical Characteristics and Laboratory Results of LAD Calcification in ESKD Patients**

| **Variables** | **LAD = 0**  **（n = 122）** | **0 < LAD ≤ 183.4**  **（n = 121）** | **LAD > 183.4**  **（n = 126）** | ***P*** |
| --- | --- | --- | --- | --- |
| **Demographics** |  |  |  |  |
| Age (years) | 44.16 ± 11.81 | 52.11 ± 11.76 | 51.90 ± 11.23 | ***< 0.001^*^*** |
| Women,n (%) | 55.00(45.08) | 52.00(42.98) | 47.00(37.30) | 0.214 |
| BMI (kg/m^2^) | 21.38 ± 3.40 | 23.16 ± 3.82 | 23.00 ± 3.95 | ***0.001^*^*** |
| SBP (mmHg) | 140.10 ± 23.33 | 137.33 ± 20.87 | 139.36 ± 24.38 | 0.806 |
| DBP (mmHg) | 87.50 ± 15.22 | 80.75 ± 14.22 | 83.43 ± 13.54 | ***0.029^*^*** |
| **Dialysis mode, n (%)** |  |  |  |  |
| Predialysis | 20.00(16.39) | 16.00(13.22) | 8.00(6.35) | ***0.014^*^*** |
| Hemodialysis | 61.00(50.00) | 77.00(63.64) | 99.00(78.57) | ***< 0.001^*^*** |
| Peritoneal dialysis | 37.00(30.33) | 29.00(24.17) | 19.00(15.20) | ***0.005^*^*** |
| **Dialysis vintage(mo)** | 60.00(3.88-97.75) | 66.00(20.00-108.00) | 96.00(51.00-132.75) | ***< 0.001^*^*** |
| **Comorbidities, n (%)** |  |  |  |  |
| Diabetic mellitus | 12.00(9.84) | 24.00(19.83) | 23.00(18.25) | 0.073 |
| Hypertension | 87.00(71.31) | 95.00(78.51) | 105.00(83.33) | ***0.023^*^*** |
| **Cause of ESKD, n (%)** |  |  |  |  |
| CGN | 91.00(74.59) | 76.00(62.81) | 88.00(69.84) | 0.430 |
| DN | 9.00(7.38) | 16.00(13.22) | 13.00(10.32) | 0.455 |
| HN | 2.00(1.64) | 5.00(4.13) | 2.00(1.59) | 0.968 |
| Polycystic kidney disease | 5.00(4.10) | 6.00(5.00) | 7.00(5.60) | 0.586 |
| Other | 15.00(12.30) | 19.00(15.7) | 16.00(12.70) | 0.933 |
| **Medication history, n (%)** |  |  |  |  |
| Lipid-lowering treatment | 10.00(8.26) | 17.00(14.05) | 16.00(12.80) | 0.274 |
| Dihydropyridine CCBs | 62.00(51.24) | 62.00(51.24) | 70.00(56.00) | 0.454 |
| ACEI/ARB | 38.00(31.40) | 28.00(23.14) | 45.00(36.29) | 0.399 |
| β-receptor blocker | 45.00(37.19) | 47.00(38.84) | 61.00(48.80) | 0.064 |
| Phosphate binders | 49.00(40.50) | 41.00(33.88) | 56.00(44.80) | 0.482 |
| Active vitamin D sterols | 42.00(34.71) | 44.00(36.36) | 45.00(36.00) | 0.835 |
| Cinacalcet | 29.00(23.97) | 32.00(26.45) | 39.00(31.20) | 0.203 |
| **Laboratory values** |  |  |  |  |
| Hemoglobin (g/l) | 98.00 ± 20.41 | 102.10 ± 19.04 | 103.96 ± 20.79 | ***0.020^*^*** |
| Hematocrit (%) | 30.15 ± 6.51 | 31.79 ± 6.11 | 32.48 ± 6.48 | ***0.004^*^*** |
| Glucose (mmol/l) | 4.41 ± 1.15 | 4.75 ± 2.22 | 4.70 ± 2.36 | 0.263 |
| Creatinine (μmol/l) | 906.40 ± 318.97 | 897.84 ± 307.34 | 877.73 ± 288.69 | 0.458 |
| Urea (mmol/l) | 23.59 ± 9.39 | 24.54 ± 12.13 | 21.71 ± 7.03 | 0.126 |
| TC (mmol/l) | 4.32 ± 1.45 | 4.20 ± 1.01 | 3.98 ± 1.11 | ***0.024^*^*** |
| TG (mmol/l) | 1.75 ± 1.26 | 1.63 ± 1.01 | 1.84 ± 1.39 | 0.547 |
| LDL-C (mmol/l) | 2.72 ± 1.08 | 2.62 ± 0.74 | 2.48 ± 0.84 | ***0.034^*^*** |
| HDL-C (mmol/l) | 1.06 ± 0.32 | 1.00 ± 0.27 | 0.92 ± 0.26 | ***< 0.001^*^*** |
| Lpa (mmol/l) | 357.93 ± 267.59 | 327.03 ± 283.54 | 313.10 ± 292.98 | 0.212 |
| Albumin (g/l) | 37.72 ± 5.52 | 36.65 ± 5.74 | 37.16 ± 4.77 | 0.422 |
| Ca (mmol/l) | 2.30 ± 0.25 | 2.33 ± 0.28 | 2.43 ± 0.24 | ***< 0.001^*^*** |
| Adjusted Ca (mmol/l) | 2.35 ± 0.23 | 2.40 ± 0.24 | 2.49 ± 0.22 | ***< 0.001^*^*** |
| Phosphorus (mmol/l) | 1.98 ± 0.45 | 2.05 ± 0.52 | 2.16 ± 0.55 | ***0.005^*^*** |
| Ca×P (mmol^2^/l^2^) | 4.65 ± 1.17 | 4.95 ± 1.40 | 5.39 ± 1.50 | ***< 0.001^*^*** |
| ALP (U/l) | 117.55(78.78-224) | 141.00(82.00-255.00) | 168.25(102.22-335.58) | 0.595 |
| Log(ALP) | 5.00 ± 0.96 | 5.12 ± 0.90 | 5.28 ± 0.88 | ***0.016^*^*** |
| BAP (μg/l) | 27.26 ± 28.32 | 31.81 ± 26.56 | 44.30 ± 38.73 | ***< 0.001^*^*** |
| Log(BAP) | 2.86 ± 1.09 | 3.17 ± 0.75 | 3.39 ± 0.92 | ***< 0.001^*^*** |
| 25-OH-D (ng/dl) | 43.57 ± 30.28 | 40.15 ± 25.90 | 46.69 ± 25.37 | 0.361 |
| iPTH (pg/ml) | 518.50(184.47-1440.33) | 856.40(239.30-1302.55) | 1203.90(358.65-1779.15) | ***0.005^*^*** |
| Log(iPTH) | 6.15 ± 1.24 | 6.34 ± 1.27 | 6.60 ± 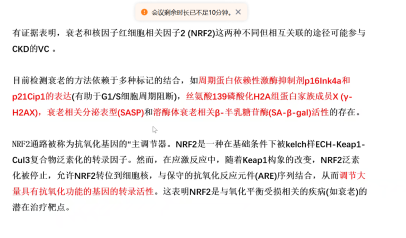1.22 | ***0.004^*^*** |

Abbreviations: ESKD = end-stage kidney disease; BMI = body mass index; SBP = systolic blood pressure; DBP = diastolic blood pressure; CGN = chronic glomerulonephritis; DN = diabetic nephropathy; HN = hypertensive nephropathy; CCB = calcium channel blocker; ACEI/ARB = angiotensin-converting enzyme inhibitors/angiotensin II receptor blockers; TC = total cholesterol; TG = triglyceride; LDL-C = low-density lipoprotein cholesterol; HDL-C = high-density lipoprotein cholesterol; Lpa = lipoprotein a; Ca = calcium; P = phosphorus; ALP = alkaline phosphatase; BAP = bone-type alkaline phosphatase; 25-OH-D = 25 hydroxyvitamin D; iPTH = intact parathyroid hormone. *P*-values are obtained from comparisons among the three groups. An asterisk(*) indicates *P* < 0.05. Data forms are expressed as mean ± standard deviation or median (Q1-Q3), except where indicated.

**Table S2. Univariate Regression Analysis of Clinical Data and Severe CACS in Patients with ESKD**

| **Variables** | **LAD CACS** |  | **Total CACS** |  |
| --- | --- | --- | --- | --- |
|  | **OR (95%CI)** | ***P*** | **OR (95%CI)** | ***P*** |
| **Demographics** |  |  |  |  |
| Age (years) | 1.027 (1.008,1.045) | ***0.005^*^*** | 1.042 (1.023,1.062) | ***< 0.001^*^*** |
| Women,n (%) | 0.756 (0.487,1.175) | 0.214 | 0.772 (0.499,1.195) | 0.246 |
| BMI (kg/m^2^ ) | 1.053 (0.993,1.116) | 0.086 | 1.027 (0.969,1.088) | 0.367 |
| SBP (mmHg) | 1.001 (0.992,1.011) | 0.800 | 0.999 (0.990,1.009) | 0.928 |
| DBP (mmHg) | 0.997 (0.982,1.012) | 0.656 | 0.988 (0.974,1.003) | 0.113 |
| **Dialysis mode, n (%)** |  |  |  |  |
| Predialysis | 0.390 (0.175,0.867) | ***0.021^*^*** | 0.370 (0.166,0.822) | ***0.015^*^*** |
| Hemodialysis | 2.790 (1.700,4.579) | ***< 0.001^*^*** | 2.989 (1.823,4.903) | ***< 0.001^*^*** |
| Peritoneal dialysis | 0.478 (0.272,0.840) | ***0.010^*^*** | 0.444 (0.253,0.779) | ***0.005^*^*** |
| **Dialysis vintage(mo)** | 1.007 (1.003,1.011) | ***< 0.001^*^*** | 1.008 (1.004,1.011) | ***< 0.001^*^*** |
| **Comorbidities, n (%)** |  |  |  |  |
| Diabetic mellitus | 1.284 (0.723,2.280) | 0.393 | 1.320 (0.746,2.334) | 0.340 |
| Hypertension | 1.676 (0.966,2.908) | 0.066 | 2.276 (1.282,4.041) | ***0.005^*^*** |
| **Cause of ESKD, n (%)** |  |  |  |  |
| CGN | 1.054 (0.661,1.682) | 0.826 | 1.009 (0.635,1.603) | 0.969 |
| DN | 1.003 (0.494,2.036) | 0.993 | 1.081 (0.539,2.170) | 0.826 |
| HN | 0.544 (0.111,2.657) | 0.452 | 0.518 (0.106,2.530) | 0.416 |
| Polycystic kidney disease | 1.246 (0.471,3.297) | 0.658 | 1.527 (0.587,3.970) | 0.386 |
| Other | 0.894 (0.473,1.691) | 0.731 | 0.939 (0.501,1.761) | 0.845 |
| **Medication history, n (%)** |  |  |  |  |
| Lipid-lowering treatment | 1.169 (0.604,2.261) | 0.643 | 1.238 (0.644,2.377) | 0.522 |
| Dihydropyridine CCBs | 1.211 (0.785,1.869) | 0.387 | 1.323 (0.859,2.037) | 0.204 |
| ACEI/ARB | 1.519 (0.956,2.413) | 0.077 | 1.493 (0.943,2.365) | 0.088 |
| β-receptor blocker | 1.554 (1.005,2.404) | ***0.048^*^*** | 1.648 (1.068,2.543) | ***0.029^*^*** |
| Phosphate binders | 1.371 (0.884,2.125) | 0.159 | 1.697 (1.097,2.623) | ***0.017^*^*** |
| Active vitamin D sterols | 1.020 (0.651,1.601) | 0.930 | 0.808 (0.514,1.270) | 0.356 |
| Cinacalcet | 1.346 (0.835,2.168) | 0.222 | 1.414 (0.881,2.269) | 0.152 |
| **Laboratory values** |  |  |  |  |
| Hemoglobin (g/l) | 1.010 (0.999,1.021) | 0.078 | 1.009 (0.999,1.020) | 0.096 |
| Hematocrit (%) | 1.038 (1.003,1.074) | ***0.034^*^*** | 1.035 (1.001,1.071) | ***0.047^*^*** |
| Glucose (mmol/l) | 1.029 (0.926,1.143) | 0.595 | 1.031 (0.929,1.145) | 0.567 |
| Creatinine (μmol/l) | 0.999 (0.999,1.000) | 0.465 | 1.000 (0.999,1.001) | 0.745 |
| Urea (mmol/l) | 0.967 (0.943,0.996) | ***0.027^*^*** | 0.974 (0.948,1.000) | ***0.049^*^*** |
| TC (mmol/l) | 0.812 (0.671,0.984) | ***0.034^*^*** | 0.757 (0.622,0.921) | ***0.005^*^*** |
| TG (mmol/l) | 1.100 (0.928,1.303) | 0.273 | 1.012 (0.852,1.202) | 0.894 |
| LDL-C (mmol/l) | 0.775 (0.600,1.001) | 0.050 | 0.721 (0.556,0.935) | ***0.014^*^*** |
| HDL-C (mmol/l) | 0.219 (0.092,0.525) | ***< 0.001^*^*** | 0.206 (0.086,0.493) | ***< 0.001^*^*** |
| Lpa (mmol/l) | 0.999 (0.999,1.000) | 0.342 | 0.999 (0.999,1.000) | 0.127 |
| Albumin (g/l) | 0.999 (0.960,1.040) | 0.963 | 0.983 (0.944,1.023) | 0.390 |
| Ca (mmol/l) | 6.085 (2.491,14.864) | ***< 0.001^*^*** | 4.806 (2.010,11.490) | ***< 0.001^*^*** |
| Adjusted Ca (mmol/l) | 9.675 (3.458,27.070) | ***< 0.001^*^*** | 8.714 (3.160,24.031) | ***< 0.001^*^*** |
| Phosphorus (mmol/l) | 1.718 (1.125,2.626) | ***0.012^*^*** | 1.740 (1.141,2.653) | ***0.010^*^*** |
| Ca×P (mmol^2^/l^2^) | 1.361 (1.158,1.600) | ***< 0.001^*^*** | 1.356 (1.155,1.592) | ***< 0.001^*^*** |
| ALP (U/l) | 1.000 (1.000,1.001) | 0.699 | 1.000 (1.000,1.001) | 0.173 |
| Log(ALP) | 1.296 (1.027,1.636) | ***0.029^*^*** | 1.466 (1.159,1.853) | ***0.001^*^*** |
| BAP (μg/l) | 1.014 (1.005,1.022) | ***0.001^*^*** | 1.015 (1.007,1.023) | ***< 0.001^*^*** |
| Log(BAP) | 1.602 (1.170,2.193) | ***0.003^*^*** | 1.688 (1.227,2.322) | ***0.001^*^*** |
| 25-OH-D (ng/dl) | 1.006 (0.999,1.014) | 0.113 | 1.002 (0.994,1.010) | 0.611 |
| iPTH (pg/ml) | 1.000 (1.000,1.001) | ***0.008^*^*** | 1.000 (1.000,1.001) | ***< 0.001^*^*** |
| Log(iPTH) | 1.273 (1.058,1.532) | ***0.011^*^*** | 1.338 (1.110,1.613) | ***0.002^*^*** |

Abbreviations: ESKD = end-stage kidney disease; BMI = body mass index; SBP = systolic blood pressure; DBP = diastolic blood pressure; CGN = chronic glomerulonephritis; DN = diabetic nephropathy; HN = hypertensive nephropathy; CCB = calcium channel blocker; ACEI/ARB = angiotensin-converting enzyme inhibitors/angiotensin II receptor blockers; TC = total cholesterol; TG=triglyceride; LDL-C = low-density lipoprotein cholesterol; HDL-C = high-density lipoprotein cholesterol; Lpa = lipoprotein a; Ca = calcium; P = phosphorus; ALP = alkaline phosphatase; BAP = bone-type alkaline phosphatase; 25-OH-D = 25 hydroxyvitamin D; iPTH = intact parathyroid hormone. The *P*-values in the table are obtained through univariate logistic regression analysis for each index and calcification score. The asterisk （*） indicates *P* < 0.05.

**Table S3.** **Clinical Characteristics and Laboratory Results of ESKD Patients Subgrouped by CACS in the Validation Cohort**

| **Variables** | **CACS = 0**  **(n = 22)** | **0 < CACS ≤ 400**  **(n = 51)** | **CACS > 400**  **(n = 54)** | ***P*** |
| --- | --- | --- | --- | --- |
| **Demographics** |  |  |  |  |
| Age (years) | 41.14 ± 9.72 | 51.18 ± 13.9 | 58.41 ± 13.37 | ***< 0.001^*^*** |
| Women,n (%) | 13(59.09) | 16(31.37) | 18(33.33) | 0.0868 |
| BMI (kg/m^2^ ) | 21.24 ± 3.09 | 23.25 ± 3.49 | 23.81 ± 4.39 | ***0.0157^*^*** |
| SBP (mmHg) | 129.86 ± 25.83 | 137.29 ± 26.28 | 140.47 ± 20.82 | 0.0953 |
| DBP (mmHg) | 78.91 ± 15.27 | 85.16 ± 17.03 | 81.68 ± 12.31 | 0.7936 |
| **Dialysis mode, n (%)** |  |  |  |  |
| Predialysis | 2(9.09) | 8(15.69) | 2(3.7) | 0.2129 |
| Hemodialysis | 10(45.45) | 20(39.22) | 40(74.07) | ***0.0024^*^*** |
| Peritoneal dialysis | 9(42.86) | 23(45.1) | 11(20.75) | ***0.0202^*^*** |
| **Dialysis vintage(months)** | 48(15-120) | 48(9-96) | 72(30-120) | ***0.0405^*^*** |
| **Comorbidities, n (%)** |  |  |  |  |
| Diabetic mellitus | 1(4.55) | 14(28) | 10(18.52) | 0.4238 |
| Hypertension | 16(72.73) | 43(84.31) | 44(81.48) | 0.5296 |
| **Cause of ESKD, n (%)** |  |  |  |  |
| CGN | 5(22.73) | 6(11.76) | 14(25.93) | 0.4139 |
| DN | NA(NA) | 10(19.61) | 8(14.81) | 0.2314 |
| HN | 2(9.09) | 6(11.76) | 9(16.67) | 0.3370 |
| Polycystic kidney disease | NA(NA) | 2(3.92) | 1(1.85) | 0.8469 |
| Other | 15(68.18) | 30(58.82) | 24(44.44) | ***0.0414^*^*** |
| **Medication history, n (%)** |  |  |  |  |
| Lipid-lowering treatment | 2(9.09) | 15(29.41) | 15(27.78) | 0.1703 |
| Dihydropyridine CCBs | 7(31.82) | 25(49.02) | 29(53.7) | 0.1091 |
| ACEI/ARB | 10(45.45) | 18(35.29) | 24(44.44) | 0.8264 |
| β-receptor blocker | 8(36.36) | 20(39.22) | 29(53.7) | 0.1071 |
| Phosphate binders | 12(54.55) | 22(43.14) | 25(46.3) | 0.6530 |
| Active vitamin D sterols | 8(36.36) | 17(33.33) | 19(35.19) | 0.9826 |
| Cinacalcet | 7(31.82) | 9(17.65) | 10(18.52) | 0.2893 |
| **Laboratory values** |  |  |  |  |
| Hemoglobin (g/l) | 106.09 ± 24.18 | 100.88 ± 18.32 | 102.07 ± 20.84 | 0.5719 |
| Hematocrit (%) | 33.14 ± 7.54 | 31.91 ± 5.7 | 33.73 ± 9.1 | 0.5297 |
| Glucose (mmol/l) | 4.81 ± 2.18 | 4.73 ± 1.9 | 4.42 ± 1.04 | 0.2795 |
| Creatinine (μmol/l) | 879.79 ± 398.25 | 850.69 ± 329.25 | 877.01 ± 273.98 | 0.9210 |
| Urea (mmol/l) | 19.72 ± 7.48 | 21.11 ± 7.3 | 32.34 ± 66.31 | 0.1737 |
| TC (mmol/l) | 4.24 ± 1.01 | 4.04 ± 1.17 | 4.03 ± 1.28 | 0.5304 |
| TG (mmol/l) | 1.73 ± 1.27 | 1.87 ± 1.14 | 1.66 ± 1.39 | 0.6756 |
| LDL-C (mmol/l) | 2.5 ± 0.68 | 2.5 ± 0.78 | 2.42 ± 0.81 | 0.6105 |
| HDL-C (mmol/l) | 1.14 ± 0.34 | 0.91 ± 0.29 | 0.96 ± 0.35 | 0.1018 |
| Lpa (mmol/l) | 201.39 ± 208.62 | 313.34 ± 297.11 | 299.66 ± 314.85 | 0.2945 |
| Albumin (g/l) | 37.25 ± 3.8 | 34.65 ± 6.25 | 36.99 ± 5.24 | 0.6266 |
| Ca (mmol/l) | 2.33 ± 0.25 | 2.19 ± 0.32 | 2.32 ± 0.28 | 0.5339 |
| Adjusted Ca (mmol/l) | 2.38 ± 0.26 | 2.3 ± 0.31 | 2.39 ± 0.26 | 0.6201 |
| Phosphorus (mmol/l) | 2 ± 0.62 | 1.85 ± 0.6 | 2.16 ± 0.6 | 0.1030 |
| Ca×P (mmol^2^/l^2^) | 4.78 ± 1.61 | 4.25 ± 1.44 | 5.17 ± 1.48 | 0.0750 |
| ALP (U/l) | 99(72-186.5) | 99(74.25-184.75) | 98.5(67.25-136.25) | 0.4378 |
| Log(ALP) | 4.85 ± 0.88 | 4.95 ± 1.04 | 4.77 ± 0.76 | 0.5730 |
| BAP (μg/l) | 59.24 ± 35.27 | 30.72 ± 37.56 | 26.23 ± 20.27 | ***0.0227^*^*** |
| Log(BAP) | 3.92 ± 0.62 | 3 ± 0.85 | 3.03 ± 0.68 | ***0.0365^*^*** |
| 25-OH-D (ng/dl) | 272.85 ± 1078.77 | 48.9 ± 101.18 | 37.9 ± 24.53 | 0.0852 |
| iPTH (pg/ml) | 410.65(70.5-858.78) | 245.9(109.1-513.45) | 276.3(96.2-743.9) | 0.9329 |
| Log(iPTH) | 5.55 ± 1.46 | 5.41 ± 1.61 | 5.38 ± 1.75 | 0.7086 |

Abbreviations: ESKD = end-stage kidney disease; BMI = body mass index; SBP = systolic blood pressure; DBP = diastolic blood pressure; CGN = chronic glomerulonephritis; DN = diabetic nephropathy; HN = hypertensive nephropathy; CCB = calcium channel blocker; ACEI/ARB = angiotensin-converting enzyme inhibitors/angiotensin II receptor blockers; TC = total cholesterol; TG = triglyceride; LDL-C = low-density lipoprotein cholesterol; HDL-C = high-density lipoprotein cholesterol; Lpa = lipoprotein a; Ca = calcium; P = phosphorus; ALP = alkaline phosphatase; BAP = bone-type alkaline phosphatase; 25-OH-D = 25 hydroxyvitamin D; iPTH = intact parathyroid hormone. The *P*-values in the table are obtained through univariate logistic regression analysis of each index and calcification score. An asterisk (*) indicates a *P-*value less than 0.05.
